# Genome-wide association study of ACE inhibitor-induced cough implicates neuropeptides and shows genetic overlap with chronic dry cough

**DOI:** 10.1101/2022.06.30.22277097

**Authors:** Kayesha Coley, David J. Shepherd, Richard Packer, Catherine John, Robert C. Free, Edward J. Hollox, Louise V. Wain, Martin D. Tobin, Chiara Batini

**Author notes:** Correspondence (KC) and (CB).

## Abstract

ACE inhibitors (ACEIs) are commonly prescribed for hypertension, a global risk factor for cardiovascular disease. Their primary side effect is a dry cough which affects 5-35% of users. As clinical guidelines recommend switching those experiencing cough to an angiotensin-II receptor blocker, we have used this switch as a proxy for ACEI-induced cough. Through a two-stage multi-ancestry genome-wide association study, including up to 7,030 cases and 39,921 controls, we identify five independent genome-wide significant associations implicating six protein-coding genes, including *INHBC, KCNIP4, NTSR1* and *PREP* which encode proteins involved in the nervous system. We also observe genetic overlap between ACEI-induced cough and chronic dry cough through genetic correlation and phenome-wide association studies. In line with existing hypotheses, our findings suggest a neurological basis for the pathology of ACEI-induced cough, particularly the role of proinflammatory mediators in sensory airway sensitivity and cough reflex modulation, and shared biological mechanisms with chronic dry cough.

## Introduction

Angiotensin-converting enzyme inhibitors (ACEIs) are a class of medications mainly used to treat hypertension and heart failure^1^, and are among the most commonly prescribed drugs^2,3^.

ACEIs reduce blood pressure by inhibiting the activity of ACE, an important component of the renin-angiotensin-aldosterone system (RAAS). ACE inhibition blocks the conversion of angiotensin I to angiotensin II and the degradation of bradykinin and substance P, thus preventing the constriction of blood vessels^4^.

Although ACEIs are generally well tolerated, the most common side effect of their use is a dry (non-productive) persistent cough, which affects 5-35%^5^ of users. Gender and ethnicity are known risk factors associated with this side effect: cough is 1.5 times more likely in females compared with males^6^, and 2.5 times more likely in East Asian populations than white populations^7^. ACEI-induced cough can only be treated by discontinuing treatment^5^ and current prescribing guidelines recommend switching to an angiotensin-II receptor blocker (ARB)^8^, a better-tolerated anti-hypertensive^9^. As a result, switching from an ACEI to an ARB is a marker for ACEI-induced cough^10^ that can be utilised in genetic studies^11^.

The mechanisms underlying ACEI-induced cough have been poorly understood. Early hypotheses have suggested this is a response to the accumulation of inflammatory neuropeptides, particularly bradykinin and substance P, upon ACE inhibition^5,12^. Genes involved in these pathways have formed the basis of many candidate gene studies^13–20^, but these have failed to yield replicable findings and proved inconclusive about the role of these mediators in ACEI-induced cough. In light of this, several genome-wide association studies^11,21,22^ (GWASs) have since been performed, the largest of which included 1,595 cases and 5,485 controls from the Electronic Medical Records and Genomics (eMERGE) Network and identified a genome-wide significant signal within an intronic region of *KCNIP4*, which was replicated in two independent datasets^11^. However, larger sample sizes are likely to be required to discover additional genetic associations, improve the fine-mapping of associated loci and understand the biological pathways involved in ACEI-induced cough.

Here, we have used a class switch from an ACEI to an ARB in electronic health records (EHRs) to define a proxy phenotype of ACEI-induced cough, and performed a multi-ancestry GWAS including over three times the number of cases studied to date. We map associated variants to putative causal genes and by studying shared genetic architecture and conducting phenome-wide association studies (PheWASs), provide insights into the shared aetiology of ACEI-induced cough and chronic dry cough.

## Results

### Study-level characteristics

Using primary care EHRs linked to UK Biobank^23^ and the Extended Cohort for E-health, Environment and DNA (EXCEED)^24^ study, cases were defined as individuals who switched from an ACEI to an ARB within 12 months of initiating ACEI treatment (*index date*) and were assumed to have switched due to cough, while controls were defined as continuous users of ACEIs. A description of the characteristics of cases and controls in both cohorts is presented in **Table 1**. The overall prevalence of ACEI-induced cough was 13.6% and 15.2% in UK Biobank and EXCEED, respectively; however, stratifying by genetically defined ancestry showed a higher prevalence in individuals of East Asian (21.6% vs 13.5%, *p*-value = 0.004) and South Asian (16.2% vs 13.5%, *p*-value = 0.011) ancestries, compared to Europeans in UK Biobank (**Table 1**). Supporting the use of the drug class switch as a proxy for ACEI-induced cough, the number of clinical cough codes in 12 months after *index date* was also associated with case status (*p*-value < 0.001; **Supplementary Table 1**).

**Table 1.**
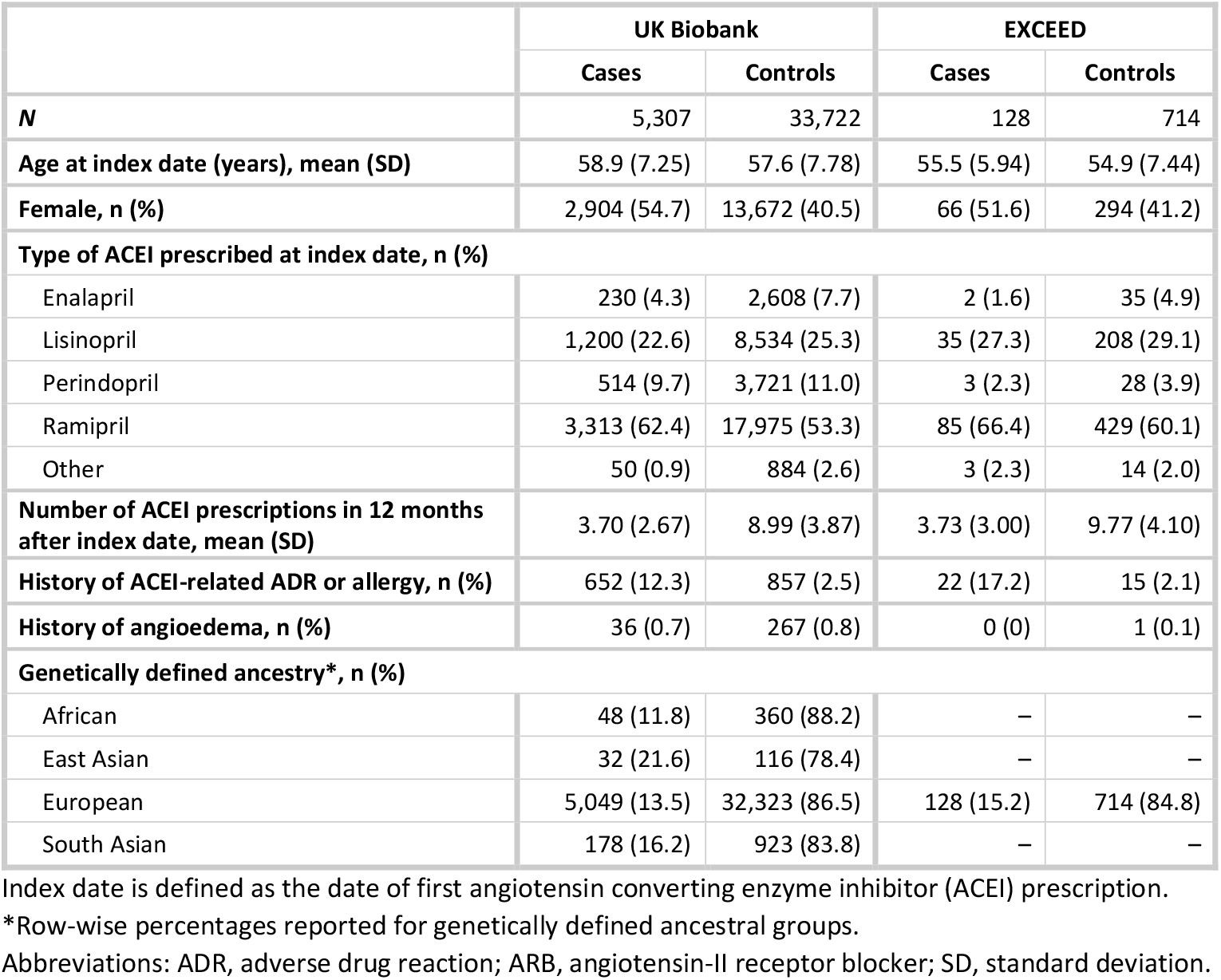
Study-level characteristics of Stage 1 meta-analysis cohorts.

### Discovery of associated genomic loci

A two-stage multi-ancestry meta-analysis approach was used to identify genome-wide significant signals associated with ACEI-induced cough (**Supplementary Figure 1**). A total of 5,435 cases and 34,436 controls from four ancestral groups in UK Biobank (African, East Asian, European and South Asian) and one ancestral group from EXCEED (European) were included in the Stage 1 meta-analysis of ∼17M variants (**Figure 1; Supplementary Figure 2**; **Supplementary Table 2**). The Stage 1 meta-analysis had a genomic inflation factor (λ_GC_) of 0.998, hence no post meta-analysis genomic control correction was performed. From the variants reaching genome-wide significance (*p*-value < 5×10^−8^), we identified 9 sentinel variants; following conditional analysis, we found one additional independent variant (rs78598167) reaching genome-wide significance, resulting in 10 signals across 9 loci which were followed up in the eMERGE Network study^11^ (Stage 2). Of these, five were genome-wide significant in the joint meta-analysis of Stage 1 and Stage 2 (**Table 2A**), three were genome-wide significant in Stage 1, but were not represented in Stage 2 (**Table 2B**), and the remaining two did not reach genome-wide significance in the joint meta-analysis (**Supplementary Table 3**). Overall, we found 8 genome-wide significant signals (5 in the joint meta-analysis, and 3 in Stage 1 only) located across 7 loci. One of the signals (rs1495509) corresponded to the previously described association in *KCNIP4*^11^, and the remainder were novel.

**Figure 1.**
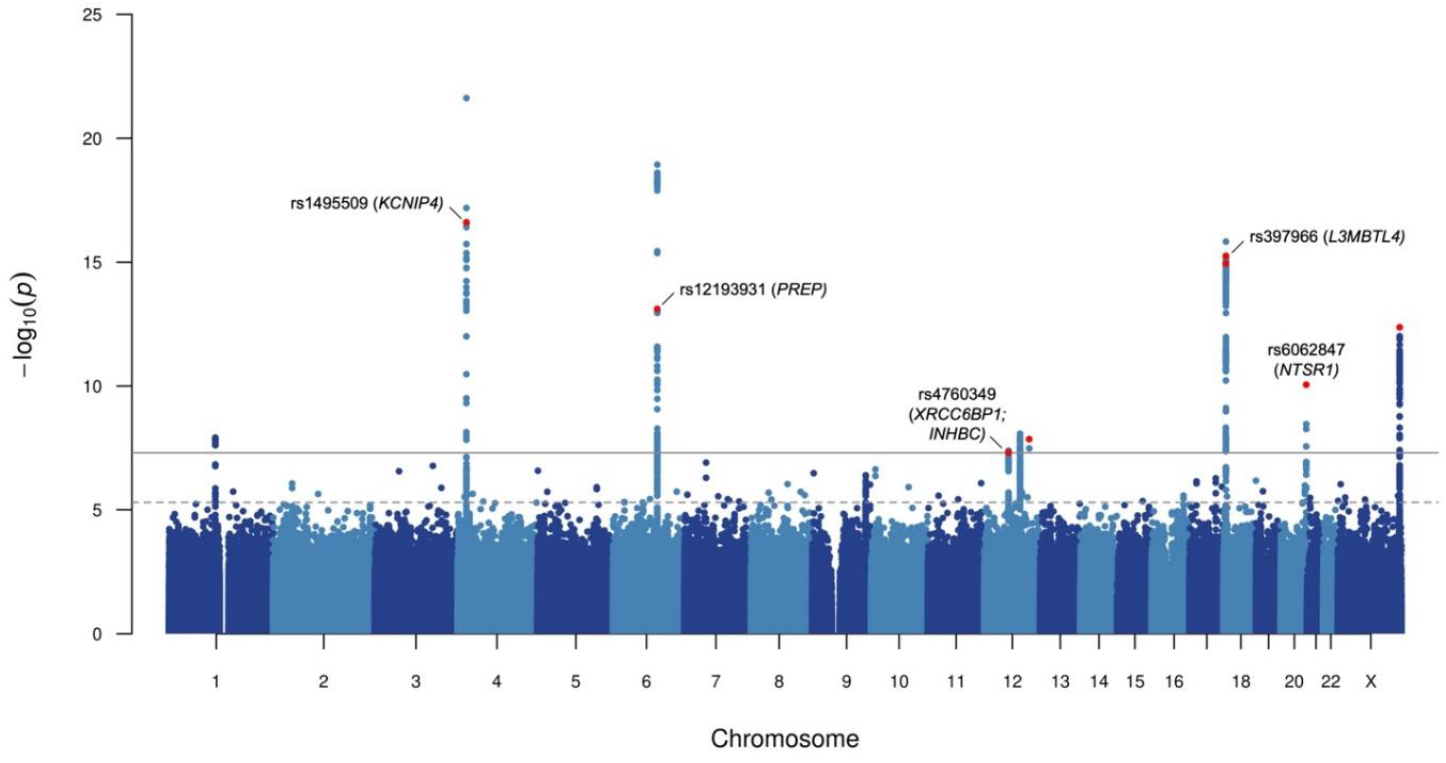
Manhattan plot for Stage 1 genome-wide meta-analysis of ACEI-induced cough. Each point represents a genetic variant, with the x-axis relating to genomic position (build hg19) and y-axis relating to –log_10_(*p*-value) of association. The dotted grey horizontal line represents the suggestive significance threshold (*p*-value of 5×10^−6^) and the solid grey horizontal line represents the genome-wide significance threshold (*p*-value of 5×10^−8^). Variants highlighted in red were either genome-wide significant in joint meta-analysis of Stage 1 and Stage 2, or genome-wide significant in Stage 1 only (without representation in Stage 2). Variants reaching genome-wide significance in joint meta-analysis are also annotated with rs number and mapped genes identified through fine-mapping of respective loci.

**Table 2.**
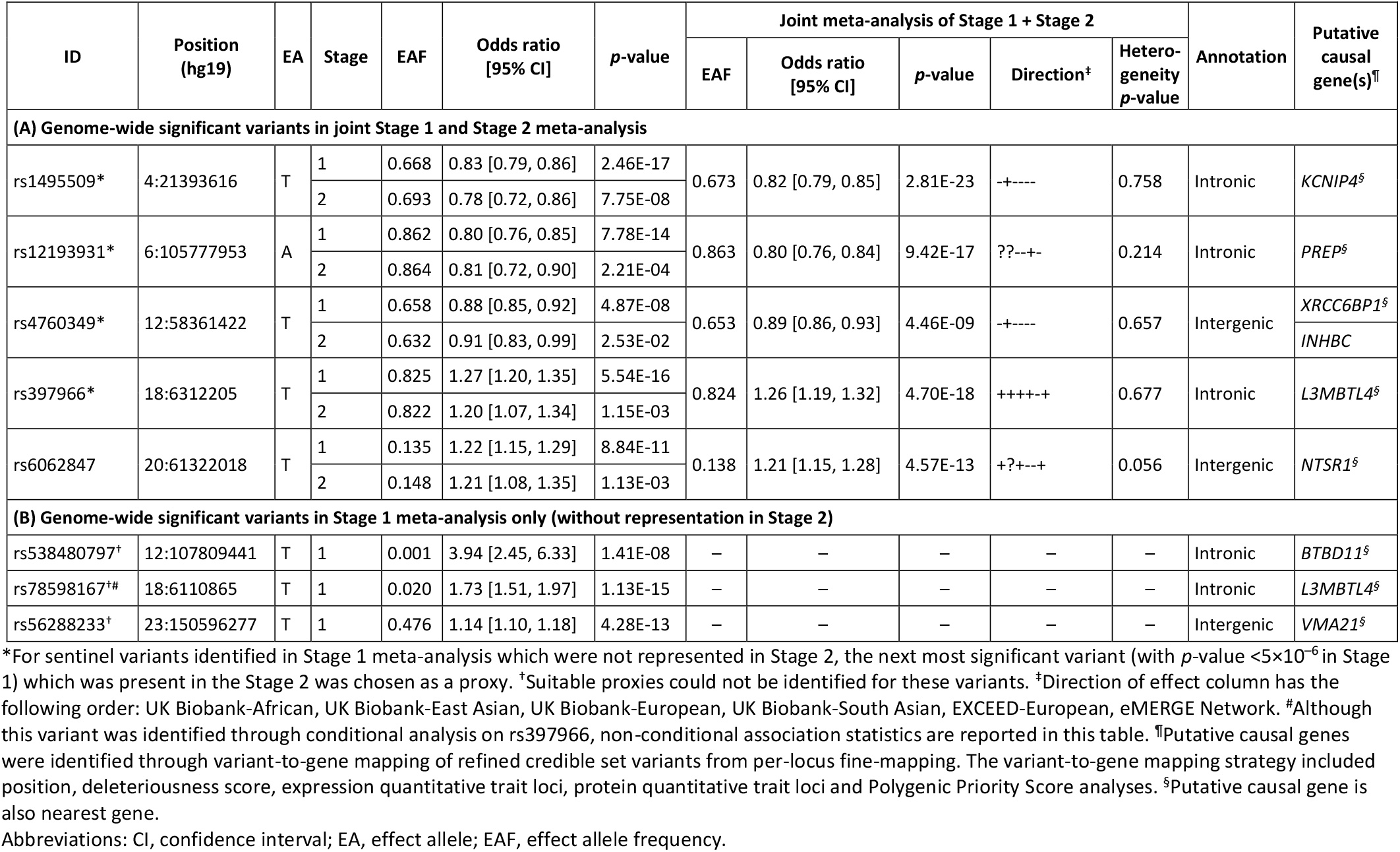
Independent sentinel variants associated with ACEI-induced cough identified from genome-wide meta-analysis of multiple ancestral groups in UK Biobank and EXCEED (Stage 1), corresponding results in the eMERGE Network (Stage 2), and joint analysis of both stages. Stage 1 included a maximum of 5,435 cases and 34,436 controls and Stage 2 included a maximum of 1,595 cases and 5,485 controls.

### Identification of putative causal variants and genes

Through fine-mapping of these 7 loci, we obtained 8 credible sets relating to each of the signals in **Table 2**, which included between 1 and 51 variants. Four out of the 148 credible set variants had a posterior inclusion probability [PIP] >50%. We defined refined credible sets containing a total of 17 variants with PIP ≥10%. All of these were annotated as non-coding (**Supplementary Table 4**) and were used as the input for our variant-to-gene mapping strategy to identify putative causal genes involved in the biological mechanism of ACEI-induced cough. The list of genes identified and the refined credible set variants supporting the mapping for each criterion are described in **Supplementary Table 5**.

Firstly, mapping based on position identified 7 unique nearest genes, and secondly CADD annotation of non-coding variants implicated an additional two genes (**Supplementary Table 4**). Thirdly, expression quantitative trait loci (eQTL) data were used to identify a total of 9 variants across 5 credible sets as significantly associated with the expression of 15 genes (FDR <0.05) in at least one of the 34 relevant tissues from the GTEx v8^25^, eQTLGen^26^ and BRAINEAC^27^ reference datasets (**Supplementary Figure 3**; **Supplementary Table 6**). Fourth, we used protein quantitative trait loci (pQTL) data to identify 5 variants across two credible sets significantly affecting (*p*-value <1.8×10^−9^) the levels of 5 proteins (ARHGEF25, INHBC, LRIG3, PREP in *cis* and DCUN1D1 in *trans*; **Supplementary Table 7**) in blood plasma using the deCODE^28^ dataset (but none in the SCALLOP Consortium^29^ dataset). Finally, Polygenic Priority Score (PoPS)^30^-based mapping identified 7 unique genes, of which 4 were also defined as the nearest gene (**Supplementary Table 8**).

Combining all five elements of our variant-to-gene mapping strategy, 23 unique protein-coding genes were identified (**Supplementary Figure 4**), of which 8 were supported by at least two criteria (**Figure 2**). The 8 putative causal genes are *BTBD11, INHBC, KCNIP4, L3MBTL4, NTSR1, PREP, VMA21* and *XRCC6BP1*, and their biological functions were explored through interrogating online databases^31–33^ and reviewing published literature (**Supplementary Table 9**). Prolyl Endopeptidase (PREP) is part of the RAAS, a physiological system which regulates multiple biological processes, including blood pressure and inflammation^34^, through the activity of two key enzymes, ACE and its homolog ACE2, which have opposing effects^35^. ACE converts angiotensin I to angiotensin II, while ACE2 degrades angiotensin I into angiotensin (1-7)^35^. PREP functions, alongside ACE2, as an angiotensin hydrolysing enzyme, and also metabolises other vasoactive peptides such as bradykinin^36,37^. Neurotensin Receptor 1 (NTSR1) is a mediator of the peripheral functions of the peptide hormone and central nervous system neurotransmitter, neurotensin^38^ which is also hydrolysed by ACE2^36,38^. Additionally, NTSR1 is a positive regulator of γ-aminobutyric acid (GABA) secretion^32^. GABA is one of the main inhibitory neurotransmitters within the central nervous system, and has additional peripheral roles including anti-inflammation^39^. L3MBTL Histone Methyl-Lysine Binding Protein 4 (L3MBTL4) and Inhibin Subunit Beta C (INHBC) are both involved in the metabolism of glycoprotein hormones. L3MBTL4 also negatively regulates transcription through chromatin organisation and histone binding activity^31^ and, following evidence of association between *L3MBTL4* and hypertension, *in vivo* analyses demonstrated that L3MBTL4 induces the proliferation and remodelling of vascular smooth muscle cells via MAPK pathway components, p38 and JNK, which results in hypertension^40,41^. As a member of the transforming growth factor (TGF)-beta protein superfamily, INHBC also has a role in SMAD protein signal transduction, and its expression in sensory neurons is required to suppress inflammatory pain^42^. Similarly, BTB Domain Containing 11 (BTBD11) is involved in SMAD protein signal transduction, and has been identified as an inhibitory interneuron-specific protein localised to the brain cortex and hippocampus and interacts with glutamatergic synapses^43^. Vacuolar ATPase assembly factor VMA21 (VMA21) is a chaperone for the assembly of lysosomal vascular ATPase^31^, and XRCC6 Binding Protein 1 (XRCC6BP1) is a metallopeptidase also involved in ATP synthase assembly, as well as DNA repair^31^. Beyond our novel findings, *KCNIP4* (Potassium Voltage-Gated Channel Interacting Protein 4) is predominantly expressed in the brain^32^ and encodes an integral component of Kv4 channel complexes. These complexes have a role in the regulation of potassium ion transmembrane transport and neuronal excitability in response to changes in calcium concentrations^31^.

**Figure 2.**
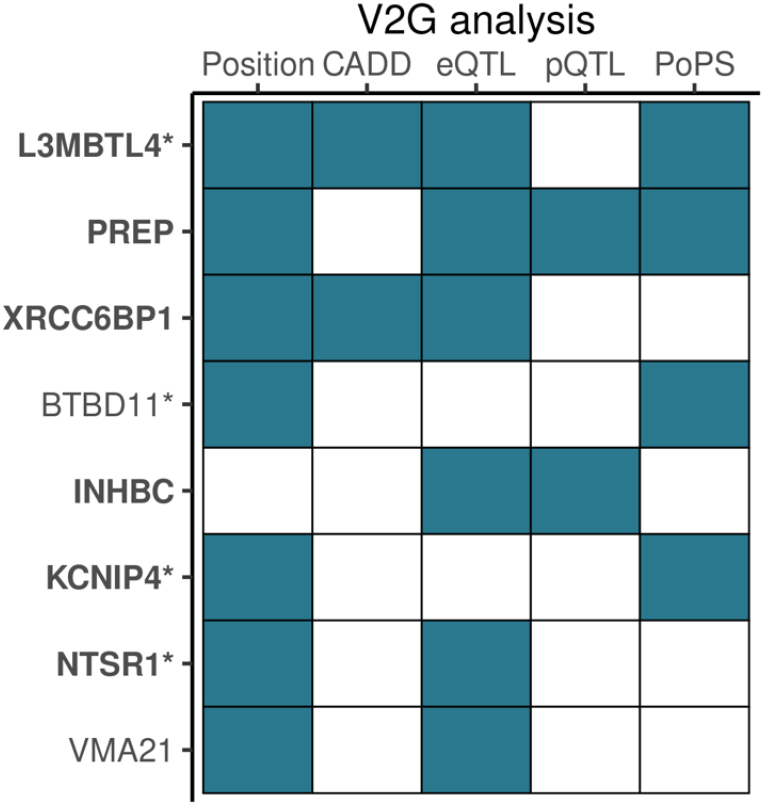
Genes identified by at least two variant-to-gene (V2G) mapping analyses based on 95% credible set variants with PIP >10% (refined credible set). Filled tiles denote gene identified by specific analysis. Genes identified by at least one refined credible set variant from fine-mapping of genome-wide significant loci in joint meta-analysis are highlighted bold, and nonbold if the fine-mapped locus was genome-wide significant in Stage 1 only (without representation in Stage 2). ^*^Genes identified by at least one mapping analysis with a refined credible set variant with PIP >50%. Abbreviations: CADD, Combined Annotation Dependent Depletion; eQTL, expression quantitative trait loci; PoPS, Polygenic Priority Score; pQTL, protein quantitative trait loci.

### Genetic architecture, overlap and pleiotropic effects of associated variants

For each variant with the highest PIP per credible set, we performed a PheWAS of up to 1,913 UK Biobank-derived phenotypes using Deep-PheWAS^44^. Three out of the 8 variants were associated (false discovery rate [FDR] ≤0.01) with at least one trait (**Supplementary Table 10**). Aligning the effect direction to the allele conferring increased risk of ACEI-induced cough, rs6062847-T (*NTSR1*) was associated with increased risk of chronic dry cough (**Supplementary Figure 5A**), revealing a shared genetic determinant between ACEI-induced cough and chronic cough. Further, rs12827878-T (*XRCC6CP1*; *INHBC*) was associated with lower urate and urea levels (**Supplementary Figure 5B**), and rs56288233-T (*VMA21*) was associated with lower insulin-like growth factor 1 levels (**Supplementary Figure 5C**).

Utilising published GWASs results, we observed that the highest PIP variants per credible set also have genome-wide significant associations across other traits (**Supplementary Table 11**). Focussing on the novel findings, we found rs878033 (*L3MBTL4*) was in linkage disequilibrium (LD; *r*^*2*^ = 1.0) with rs403814-A which increased risk of both ACEI-induced cough and hypertension^40^.

Using LD-score regression (LDSC)^45,46^ and the UK Biobank European-only ACEI-induced cough analysis, we generated estimates of narrow sense SNP-heritability (*h*^*2*^) and calculated genetic correlation (*r*_*g*_) with chronic dry cough. The estimated *h*^*2*^ of ACEI-induced cough was 5.1% (SE 1.4%) and we found strong genetic correlation between ACEI-induced cough and chronic dry cough (*r*_*g*_ = 0.56 [SE 0.15], *Z*-score = 3.78, *p*-value = 0.0002), further supporting genetic overlap with this phenotype.

## Discussion

We present the largest multi-ancestry pharmacogenomic study of ACEI-induced cough to date, implicating genes involved in the nervous system and neurogenic inflammation and highlighting genetic overlap between ACEI-induced cough and chronic dry cough.

In a two-stage meta-analysis (UK Biobank^23^, EXCEED^24^ and eMERGE Network^11^), we identified 5 genome-wide significant sentinels in the joint Stage 1 and Stage 2 meta-analysis which were mapped to 6 protein-coding genes (*INHBC, KCNIP4, L3MBTL4, NTSR1, PREP* and *XRCC6BP1*), and a further 3 genome-wide significant sentinels from the Stage 1 analysis only, which were mapped to 2 additional genes (*BTBD11* and *VMA21*). Only one of these 8 signals was previously described^11^ (rs1495509; *KCNIP4*), resulting in 7 novel signals and 7 novel mapped genes. The mapped genes are involved in biological processes including neuropeptide signalling, neuronal excitability, SMAD protein signal transduction, DNA repair, and regulation of transcription and ATP-dependent activity. We also observed a strong genetic correlation with chronic dry cough and a PheWAS of rs6062847-T, which mapped to *NTSR1*, showed significant associations which chronic cough phenotypes, highlighting shared genetic determinants between these traits.

We find five genes which encode proteins with important neurological functions. PREP is involved in the mediation of neuropeptide activity, including the metabolism of bradykinin^36,37^. Bradykinin is known to target airway C-fibres, and has been shown to sensitise airway sensory nerves and induce cough reflex hypersensitivity in an animal model^47,48^. Beyond the implication of NTSR1 in the mediation of proinflammatory peptide, neurotensin, NTSR1 is also known to be involved in airway smooth muscle contraction^49,50^. It is also a positive regulator of the secretion of GABA, an inhibitory neurotransmitter with anti-inflammatory activity^32^. Animal models^51,52^ and controlled trials^53,54^ have demonstrated that agonists to GABA receptors inhibit the cough reflex; specifically, baclofen has been shown to supress ACEI-induced cough^55^. Further, INHBC is expressed in the nervous system and is known to suppress inflammatory pain responses^42^ and current evidence demonstrates the role of BTBD11 in regulating glutamatergic synapse function^43^. Also, combined with additional support from previous genomic studies^11^, *in vitro* and *in vivo* studies^56^ of KCNIP4 activity provide evidence for the role of neuronal potassium channels in smooth muscle reactivity and cough reflex modulation, and also suggest that they are a promising target for antitussive therapies^57^. In summary, we provide supporting evidence for the most widely accepted hypothesis of this adverse drug reaction (ADR), which suggests the mechanism is due to bradykinin-induced sensitivity of airway sensory nerves following the accumulation of bradykinin upon ACE inhibition^5,12^. We also highlight the roles of other inflammatory mediators, including neurotensin and GABA, as well as regulators of neuronal excitability, in airway sensitivity and cough reflex modulation.

In this study, we have used EHRs to define a proxy phenotype for ACEI-induced cough using an ACEI to ARB switch. In the eMERGE Network GWAS^11^ (Stage 2), ACEI-induced cough was defined using the EHR allergy section. Although this was a more direct phenotyping approach using a confirmed diagnosis of cough as a reaction to an ACEI, we replicated the *KCNIP4* signal identified at genome-wide significance, with very comparable effect sizes, even when considering Stage 1 only; this provides a good indication that the proxy phenotype appropriately captured ACEI-induced cough. Our genome-wide discovery (Stage 1) included more than three times the number of cases than the largest previous study^11^. This powerful design allowed us to identify 7 additional variant-phenotype associations, 4 of which independently replicated in Stage 2.

We did not replicate any of the genome-wide significant variants identified in a different published GWAS study of ACEI intolerance^21^ at a Bonferroni-corrected *p*-value threshold. Although our phenotyping strategy was aligned to that of Mahmoudpour *et al*^21^, the associated variants, which implicated genes including *GABRG2, MBOAT1* and *RBFOX3*, were from a discovery-only analysis which was likely underpowered, with almost six times fewer cases than our Stage 1 analysis.

Pharmacogenomic studies are often limited by small sample sizes^58^ which presents a challenge for the discovery of genomic loci associated with pharmacological traits. Since data on drug response and ADR-related traits are rarely specifically recorded in study populations, we have harnessed the large-scale resource of EHRs to characterise ACEI-induced cough by examining drug switches from ACEI to ARB. After cough, the most probable alternative reason for a switch of this kind is angioedema. Although observed in fewer than 1% of ACEI users, angioedema is the most severe side effect and is potentially fatal^59^. However, we did not observe a significant difference in angioedema diagnoses between cases and controls, and its overall rare prevalence makes it unlikely to have an effect on genome-wide association statistics.

We also note limitations of this study. We acknowledge that EHR data are not optimised for research and may contain sources of error, incompleteness and bias^60^. Any resulting misclassification of cases and controls will tend to reduce statistical power for discovery of genetic associations. In UK Biobank we observed that only approximately 12% of individuals defined as cases had a clinical coded record of an ACEI-related ADR or allergy, consistent with previous findings from UK clinical practice^61^. In this context, ADR definitions which are not susceptible to clinician-dependent sources of error^62^ are valuable, such as the coded prescription data we utilised as an ADR indicator. The sample sizes of non-European ancestry cases and controls available to us were modest, and the findings were largely driven by the European ancestry participants. European-bias presents as an important issue for many GWASs^63^ and highlights the need for the recruitment of ancestrally diverse individuals to longitudinal population studies with linked EHRs.

In conclusion, using our EHR-based phenotype definition, we have performed the largest pharmacogenomic analysis of ACEI-induced cough to date, and identified novel putative causal variants and genes. We have provided genetic evidence of a shared pathophysiological mechanism between ACEI-induced cough and chronic dry cough, which we have shown to have a neurological basis involving inflammatory factors. Aside from providing mechanistic insights and highlighting potential shared drug targets, we have also identified candidates for predictive variants for future pharmacogenomic-guided prescribing practices to avoid reactive drug switching upon occurrence of an ADR. This has the potential to enhance drug safety and efficacy and improve patient outcomes.

## Methods

### Study populations

UK Biobank is a large longitudinal study in the UK, described extensively elsewhere^23^. With relevance to the present study, roughly 230,000 participants have linked primary care EHRs as well as genomic data on over 90 million variants through imputation to Haplotype Reference Consortium^64^ and UK10K^65^ + 1000 Genomes Project^66^.

The EXCEED study^24^ is a population-based cohort study with participants recruited from Leicester, Leicestershire and Rutland. Approximately 5,000 participants have both EHR linkage and TopMed^67^-imputed genomic data for over 300 million variants.

Our two-stage meta-analysis study design (**Supplementary Figure 1**) included UK Biobank and EXCEED in Stage 1, and genome-wide summary statistics from the eMERGE Network study^11^ in Stage 2.

### Use of EHRs for the phenotype definition and extraction of cohort characteristics

ACEI-induced cough was defined in primary care EHRs using a switch from an ACEI to an ARB as proxy phenotype. Specifically, cases were defined as individuals who switched from an ACEI to an ARB within 12 months of the date of their first ACEI prescription (defined as the *index date*) and did not receive a further ACEI prescription in the year after the switch date. Controls were defined as individuals who received at least one further ACEI prescription and no ARB prescriptions within 12 months after *index date*. Individuals were excluded if they had less than 2 years of complete data (defined as the duration of time between *index date* and either record extraction, or date of death, whichever occurred first) or received an ARB prescription before, or on the same date as, the *index date*.

ACEI and ARB prescription records were identified using a search-based strategy for UK Biobank (data providers 1,2 and 3) using a curated list of generic and brand name drugs based on the British National Formulary (BNF), dictionary of medicines and devices (dm+d) and Read v2 codes for each respective drug (**Supplementary Table 12**). For UK Biobank (data provider 4) and EXCEED, Read v2 codes were used to extract ACEI and ARB prescription events (**Supplementary Table 13**).

For descriptive analyses, clinical records for cough, ACEI-related ADR and allergy, and angioedema were extracted using the Read v2 and v3 code lists in **Supplementary Table 14**.

### Sample cohort characteristics

Statistical tests for differences between cases and controls for a number of variables were performed using either a Chi-square test of independence (categorical variables) or t-test (continuous variables).

Genetically-determined ancestry was defined using *k-*means clustering of the first two genetic principal components (PCs)^68^; for UK Biobank participants, these were provided by UK Biobank, for EXCEED participants these were calculated using EIGENSOFT. Ancestry groups were defined as African, East Asian, European and South Asian; per-ancestry sample sizes are shown in **Table 1**and **Supplementary Figure 1**.

### Genome-wide association meta-analyses

For each ancestral group in the Stage 1 studies, regenie (v1.0.6.2)^69^ was used to perform single-variant association testing. Genotyped variants with minor allele frequency (MAF) >5%, minor allele count (MAC) >100, genotype missingness <10%, Hardy-Weinberg equilibrium *p*-value >10^−15^ located outside regions of long-range LD were pruned using *--indep-pairwise 1000 100 0*.*9* in PLINK 2.0^70^ and used for model fitting. Imputed variants with imputation quality score ≥0.3, MAC_cases_ ≥10 and MAC_controls_ ≥10 were tested for association using an approximate Firth logistic regression model with age (at *index date*), age^2^, sex, genotyping array (UK Biobank only) and the first 10 genetic PCs fitted as covariates.

A two-stage meta-analysis approach (**Supplementary Figure 1**) was used for which all meta-analyses were performed using an inverse-variance weighted fixed effects model implemented in METAL^71^ with genomic control correction to account for residual population structure and relatedness. Stage 1 included all ancestry-specific summary statistics from UK Biobank and EXCEED, preserving variants represented by either (i) at least two cohort-ancestry combinations, or (ii) by only one cohort if the ancestral group was European. Stage 2 included summary statistics from a GWAS of ACEI-induced cough, defined using the allergy section of EHRs, in ancestrally diverse participants from the eMERGE Network^11^. The summary statistics from Stage 2 included 1.9M autosomal variants with an imputation quality score ≥0.7 and MAF ≥1%.

λ_GC_ was estimated as the ratio of the median of the observed Z-statistic distribution to the expected median for all individual analyses, and the Stage 1 meta-analysis, using R (v 4.1.0). Manhattan and quantile-quantile (QQ) plots were also generated using R (v 4.1.0) using the qqman package^72^.

### Signal selection and joint meta-analysis

From the Stage 1 meta-analysis, genome-wide significant variants (*p*-value <5×10^−8^) were identified and sentinels defined as the variant with the lowest *p*-value within ±1Mb regions. Where possible, sentinels which did not have representation in Stage 2 were redefined with a suitable proxy, identified as the next most significant variant within the ±1Mb region (*p*-value <5×10^−6^ in Stage 1) present in Stage 2. For each genome-wide significant locus (defined as ±1Mb from each Stage 1 sentinel variant), secondary independent association signals were identified using conditional analysis implemented in GCTA-COJO^73,74^ using *--cojo-cond* on each sentinel variant with default settings. Individual-level genomic data from UK Biobank Europeans was used as an LD reference given it was the largest contributing cohort-ancestry group in each meta-analysis. Independent sentinel variants from Stage 1 were subsequently meta-analysed with Stage 2 to generate joint meta-analysis association statistics.

Further, for fine-mapping and variant-to-gene mapping purposes, a genome-wide meta-analysis of both stages which included ∼17.2M variants represented in either Stage 1 or Stage 2 was also performed.

### Fine-mapping

Utilising summary statistics from the genome-wide meta-analysis of Stage 1 plus Stage 2, fine-mapping was performed on each locus with a genome-wide significant sentinel variant in either the joint meta-analysis, or in Stage 1 only if the sentinel variant was not represented in Stage 2.

PolyFun^75^-informed SuSiE^76^ was used to perform functionally-informed multi-variant fine-mapping of each autosomal locus. Functional prior causal probabilities specified by PolyFun were proportional to stratified LDSC (S-LDSC) heritability estimates based on the baseline-LF 2.2.UKB model, which includes variant annotations related to presence in coding regions, conservation, regulation, MAF and LD^77–79^. Pre-computed LD matrices based on unrelated British UK Biobank individuals were used as a reference. As PolyFun does not currently support X-chromosome analyses, the single-variant fine-mapping Wakefield^80^ method was used in this case, with the prior *W* set as 0.04 in the approximate Bayes factor formula. Both Bayesian methods were used to calculate 95% credible sets which were subsequently filtered for variants with PIP >10% to generate refined credible sets.

### Variant-to-gene mapping

The refined credible sets variants (and where required, LD proxies with *D’* >0.9) were used as the input for all gene mapping analyses to identify putative causal genes. The variant-to-gene mapping approach included: (i) identification of the nearest gene, (ii) variant annotation and pathogenicity estimates, (iii) eQTL analysis, (iv) pQTL analysis, and (v) PoPS.

#### (i) Nearest genes

Variants were annotated with nearest protein-coding gene within ±250kb using ANNOVAR^81^. The nearest genes were identified for positional mapping.

#### (ii) Variant annotation and pathogenicity estimates

Variants were annotated with functional consequence using ANNOVAR^81^, and with deleteriousness predictors (SIFT, PolyPhen-2 and CADD^82^) using Variant Effect Predictor^83^. The nearest genes to variants annotated as deleterious by SIFT, probably damaging or possibly damaging by PolyPhen-2 or with a CADD Phred score ≥12.37 were identified.

#### (iii) eQTL

The results of eQTL analyses from three datasets were investigated in FUMA v1.3.7^84^: GTEx v8^25^ (restricted to adrenal, artery, blood, brain, esophagus, heart, ileum, liver, lung, muscle, nerve, pituitary, spleen and stomach tissues), eQTLGen^26^ and BRAINEAC^27^. As little is known about the mechanism of ACEI-induced cough we included tissues involved in ACEI pharmacodynamics and pharmacokinetics, as well as tissues of the respiratory, cardiovascular and nervous systems. Variants with FDR <0.05 were defined as significant eQTLs. Only significant *cis*-eQTLs (i.e. within 1Mb of the gene with affected gene expression) in our refined credible sets were used to identify genes.

#### (iv) pQTL

The deCODE Genetics^28^ dataset, which includes 4,719 proteins measured by 4,907 aptamers, and the SCALLOP Consortium^29^ dataset, which includes 90 cardiovascular proteins, were investigated. To define a variant as a significant pQTL the same *p*-value thresholds were used as by the authors in the original publications, that is *p*-value <1.8×10^−9^ for deCODE Genetics and *p*-value <5×10^−8^ for the SCALLOP Consortium. Significant pQTLs were defined as *cis*-pQTLs if they were located within 1Mb of the transcription start site (TSS) of the gene encoding the measured protein or *trans*-pQTLs if they were located on a different chromosome, or more than 1Mb away from the TSS of the gene that encodes the measured protein. Only significant pQTLs in our refined credible sets were used to identify genes.

#### (v) PoPs

Finally, a similarity-based prioritisation method, PoPS^30^, which utilises genome-wide summary statistics (in this case, the genome-wide summary statistics from the Stage 1 plus Stage 2 meta-analysis) and genetic enrichments, including single-cell expression, biological pathways and protein-protein-interactions, was used to calculate gene-level associations and subsequently a priority score for all protein-coding genes. For this analysis, the European population from 1000 Genomes Project^66^ Phase 3 was used as an LD reference and the gene with the highest polygenic priority score within ±250kb of each variant was identified.

Genes supported by at least 2 out of the 5 criteria of the variant-to-gene mapping strategy were defined as putative causal genes, and database (GeneCards^31^, Open Targets Platform^32^ and Online Mendelian Inheritance in Man^33^ [OMIM]) and literature searches were used to retrieve information about their biological functionality.

### Exploring pleiotropic effects of associated variants

Further follow-up analyses were performed on each variant with the highest PIP per credible set, calculated from fine-mapping of genome-wide significant loci in joint Stage 1 and Stage 2 meta-analysis, or in Stage 1 only (without representation in Stage 2).

#### Phenome-wide association study

The Deep-PheWAS^44^ pipeline was used to perform single-variant association testing with up to 1,913 phenotypes defined in UK Biobank individuals of European ancestry. These phenotypes included: (i) composite phenotypes defined using linked hospital data, primary care data and field-IDs, (ii) Phecode-based phenotypes, (iii) field-ID phenotypes, (iv) combined field-ID phenotypes (more than two quantitative field-ID phenotypes into a single measure), all generated using the phenotype matrix generation pipeline, and (v) formula phenotypes developed within the group.

Significant variant-trait associations had a Benjamini-Hochberg corrected FDR ≤ 0.01.

#### Database queries

Open Targets Genetics^85^ was queried to identify traits for which the highest PIP variants were in LD (*r*^*2*^ >0.8) with genome-wide significant lead GWAS variants. Open Targets Genetics includes published results from NHGRI-EBI GWAS Catalogue^86^ and UK Biobank phenotypes from Neale Lab Round 2^87^ and The University of Michigan SAIGE analysis^88^.

### Genetic architecture and overlap

Utilising the UK Biobank European-only summary statistics for ACEI-induced cough, we performed two LD score regression-based analyses using LDSC^45,46^; (i) SNP-heritability, and (ii) genetic correlation with chronic dry cough, defined in UK Biobank. Both analyses utilised LD Scores pre-computed from 1000 Genomes Europeans, and all summary statistics were filtered for the HapMap3 variants.

#### Genetic correlation traits

Chronic dry cough was defined using a combination of UK Biobank field-IDs 22502 (‘cough on most days’) and 22504 (‘bring up phlegm/sputum/mucus on most days’), as in the PheWAS. The chronic dry cough GWAS included 102,941 individuals (8,388 cases; 94,553 controls) of European ancestry and was performed using regenie (v1.0.6.2)^69^ (as above) using an approximate Firth logistic regression model adjusted for age, age^2^, sex, ever smoking status, genotyping array, and the first 10 genetic principal components.

### Resource availability

#### Lead contact

Further information and requests should be directed to the lead contact, Chiara Batini.

#### Materials availability

This study did not generate new materials.

## Data Availability

UK Biobank (https://www.ukbiobank.ac.uk/enable-your-research/apply-for-access) and EXCEED (https://exceed.org.uk/research/) individual-level data are available to approved researchers upon application or data access request. GWAS summary statistics from the eMERGE Network study (Stage 2) are available from the respective study authors. GWAS summary statistics from the discovery (Stage 1) analysis are being deposited at the NHGRI-EBI GWAS Catalog (https://www.ebi.ac.uk/gwas/) and accession numbers will be added as soon as they are available. The phenotyping algorithm is publicly available (https://doi.org/10.5281/zenodo.6780065); scripts used to run additional analyses are available upon request to the lead contact.

## Acknowledgements

We thank all cohort study participants from UK Biobank and EXCEED who have made this project possible. We also thank authors of the eMERGE Network study for sharing GWAS summary statistics for use in this project. The UK Biobank genetic and phenotypic data were obtained under UK Biobank Application 59822. This study used the ALICE and SPECTRE High Performance Computing Facilities at the University of Leicester.

KC was supported by the University of Leicester (College of Life Sciences) and Health Data Research UK. CJ held a Medical Research Council Clinical Research Training Fellowship (MR/P00167X/1). LVW holds a GSK/Asthma + Lung UK Chair in Respiratory Research (C17-1). MDT holds a Wellcome Trust Investigator Award (WT202849/Z/16/Z), which has partially supported EXCEED, and NIHR Senior Investigator Award (NIHR201371). CB was supported by a UKRI Innovation Fellowship at Health Data Research UK (MR/S003762/1).

This study used data from the Extended Cohort for E-health, Environment and DNA (EXCEED) study. EXCEED is supported by the University of Leicester, the National Institute for Health and Care Research (NIHR) Leicester Respiratory Biomedical Research Centre, the Wellcome Trust (WT 202849), and Cohort Access fees from studies funded by the Medical Research Council (MRC), Biotechnology and Biological Sciences Research Council, NIHR, the UK Space Agency, and GlaxoSmithKline. It was previously supported by MRC grant G0902313. EXCEED is supported by BREATHE - The Health Data Research Hub for Respiratory Health (UKRI_PC_19004) in partnership with SAIL Databank. BREATHE is funded through the UK Research and Innovation (UKRI) Industrial Strategy Challenge Fund and delivered through Health Data Research UK. The research was partially supported by the NIHR Leicester Biomedical Research Centre and the NIHR Nottingham Biomedical Research Centre; views expressed are those of the author(s) and not necessarily those of the NHS, the NIHR or the Department of Health. This research was funded in whole, or in part, by the Wellcome Trust. This work was supported by the University of Leicester and Health Data Research UK, an initiative funded by UKRI, Department of Health and Social Care (England) and the devolved administrations, and leading medical research charities.

For the purpose of open access, the author has applied a CC BY public copyright licence to any Author Accepted Manuscript version arising from this submission.

## Author contributions

Conceptualization: MDT and CB; Methodology: KC, DJS, MDT and CB; Software: KC and RCF; Validation: KC; Formal Analysis: KC and RP; Investigation: KC, MDT and CB; Resources: DJS, RP, CJ, RCF, EJH, LVW and MDT; Data Curation: KC, CB; Writing – Original Draft: KC, MDT and CB; Writing – Review & Editing: all authors; Visualization: KC; Supervision: MDT and CB; Funding Acquisition: EJH, LVW and MDT.

## References

1. National Institute for Health and Care Excellence (2021). Hypertension: Angiotensin-converting enzyme inhibitors.

2. NHS Digital (2018). Prescriptions Dispensed in the Community - Statistics for England, 2007-2017. Prescr. Dispens. Community. https://digital.nhs.uk/data-and-information/publications/statistical/prescriptions-dispensed-in-the-community/prescriptions-dispensed-in-the-community-england---2007---2017.

3. Fuentes, A., Pineda, M., and Venkata, K. (2018). Comprehension of Top 200 Prescribed Drugs in the US as a Resource for Pharmacy Teaching, Training and Practice. Pharmacy 6, 43.

4. Herman, L.L., Padala, S.A., Ahmed, I., and Bashir, K. (2021). Angiotensin Converting Enzyme Inhibitors (ACEI). In StatPearls (StatPearls Publishing).

5. Dicpinigaitis, P. V (2006). Angiotensin-Converting Enzyme Inhibitor-Induced Cough. Chest 129, 169S–173S.

6. Alharbi, F.F., Kholod, A.A.V., Souverein, P.C., Meyboom, R.H., de Groot, M.C.H., de Boer, A., and Klungel, O.H. (2017). The impact of age and sex on the reporting of cough and angioedema with renin-angiotensin system inhibitors: a case/noncase study in VigiBase. Fundam. Clin. Pharmacol. 31, 676–684.

7. McDowell, S.E., Coleman, J.J., and Ferner, R.E. (2006). Systematic review and meta-analysis of ethnic differences in risks of adverse reactions to drugs used in cardiovascular medicine. BMJ 332, 1177–1181.

8. National Institute for Health and Care Excellence (2021). Managing angiotensin-converting enzyme inhibitors.

9. Caldeira, D., David, C., and Sampaio, C. (2012). Tolerability of Angiotensin-Receptor Blockers in Patients with Intolerance to Angiotensin-Converting Enzyme Inhibitors. Am. J. Cardiovasc. Drugs 12, 263–277.

10. Mahmoudpour, S.H., Asselbergs, F.W., de Keyser, C.E., Souverein, P.C., Hofman, A., Stricker, B.H., de Boer, A., and Maitland-van der Zee, A.H. (2015). Change in prescription pattern as a potential marker for adverse drug reactions of angiotensin converting enzyme inhibitors. Int. J. Clin. Pharm. 37, 1095–1103.

11. Mosley, J.D., Shaffer, C.M., Van Driest, S.L., Weeke, P.E., Wells, Q.S., Karnes, J.H., Velez Edwards, D.R., Wei, W.Q., Teixeira, P.L., Bastarache, L., et al. (2016). A genome-wide association study identifies variants in KCNIP4 associated with ACE inhibitor-induced cough. Pharmacogenomics J. 16, 231–237.

12. Fox, A.J., Lalloo, U.G., Belvisi, M.G., Bernareggi, M., Chung, K.F., and Barnes, P.J. (1996). Bradykinin-evoked sensitization of airway sensory nerves: a mechanism for ACE-inhibitor cough. Nat. Med. 2, 814–817.

13. Zee, R.Y.L., Rao, V.S., Paster, R.Z., Sweet, C.S., and Lindpaintner, K. (1998). Three Candidate Genes and Angiotensin-Converting Enzyme Inhibitor–Related Cough. Hypertension 31, 925–928.

14. Mukae, S., Aoki, S., Itoh, S., Iwata, T., Ueda, H., and Katagiri, T. (2000). Bradykinin B 2 Receptor Gene Polymorphism Is Associated With Angiotensin-Converting Enzyme Inhibitor–Related Cough. Hypertension 36, 127–131.

15. Mukae, S., Itoh, S., Aoki, S., Iwata, T., Nishio, K., Sato, R., and Katagiri, T. (2002). Association of polymorphisms of the renin–angiotensin system and bradykinin B2 receptor with ACE-inhibitor-related cough. J. Hum. Hypertens. 16, 857–863.

16. Kim, T.-B., Oh, S.-Y., Park, H.-K., Jeon, S.-G., Chang, Y.-S., Lee, K.-Y., Cho, Y.S., Chae, I.-H., Kim, Y.-K., Cho, S.-H., et al. (2009). Polymorphisms in the neurokinin-2 receptor gene are associated with angiotensin-converting enzyme inhibitor-induced cough. J. Clin. Pharm. Ther. 34, 457–464.

17. Grilo, A., Sáez-Rosas, M.P., Santos-Morano, J., Sánchez, E., Moreno-Rey, C., Real, L.M., Ramírez-Lorca, R., and Sáez, M.E. (2011). Identification of genetic factors associated with susceptibility to angiotensin-converting enzyme inhibitors-induced cough. Pharmacogenet. Genomics 21, 10–17.

18. Mas, S., Gassò, P., Álvarez, S., Ortiz, J., Sotoca, J.M., Francino, A., Carne, X., and Lafuente, A. (2011). Pharmacogenetic predictors of angiotensin-converting enzyme inhibitor-induced cough. Pharmacogenet. Genomics 21, 531–538.

19. Mahmoudpour, S.H., Leusink, M., Putten, L. van der, Terreehorst, I., Asselbergs, F.W., de Boer, A., and Maitland-van der Zee, A.H. (2013). Pharmacogenetics of ACE inhibitor-induced angioedema and cough: a systematic review and meta-analysis. Pharmacogenomics 14, 249–260.

20. Lee, C.J., Choi, B., Pak, H., Park, J.M., Lee, J.H., and Lee, S.H. (2022). Genetic Variants Associated with Adverse Events after Angiotensin-Converting Enzyme Inhibitor Use: Replication after GWAS-Based Discovery. Yonsei Med. J. 63, 342.

21. Mahmoudpour, S.H., Veluchamy, A., Siddiqui, M.K., Asselbergs, F.W., Souverein, P.C., De Keyser, C.E., Hofman, A., Lang, C.C., Doney, A.S.F., Stricker, B.H., et al. (2017). Meta-analysis of genome-wide association studies on the intolerance of angiotensin-converting enzyme inhibitors. Pharmacogenet. Genomics 27, 112–119.

22. Hallberg, P., Persson, M., Axelsson, T., Cavalli, M., Norling, P., Johansson, H.-E., Yue, Q.-Y., Magnusson, P.K., Wadelius, C., Eriksson, N., et al. (2017). Genetic variants associated with angiotensin-converting enzyme inhibitor-induced cough: a genome-wide association study in a Swedish population. Pharmacogenomics 18, 201–213.

23. Bycroft, C., Freeman, C., Petkova, D., Band, G., Elliott, L.T., Sharp, K., Motyer, A., Vukcevic, D., Delaneau, O., O’Connell, J., et al. (2018). The UK Biobank resource with deep phenotyping and genomic data. Nature 562, 203–209.

24. John, C., Reeve, N.F., Free, R.C., Williams, A.T., Ntalla, I., Farmaki, A.E., Bethea, J., Barton, L.M., Shrine, N., Batini, C., et al. (2019). Cohort Profile: Extended Cohort for E-health, Environment and DNA (EXCEED). Int. J. Epidemiol. 48, 1734.

25. The GTEx Consortium (2020). The GTEx Consortium atlas of genetic regulatory effects across human tissues. Science 369, 1318–1330.

26. Võsa, U., Claringbould, A., Westra, H.J., Bonder, M.J., Deelen, P., Zeng, B., Kirsten, H., Saha, A., Kreuzhuber, R., Yazar, S., et al. (2021). Large-scale cis- and trans-eQTL analyses identify thousands of genetic loci and polygenic scores that regulate blood gene expression. Nat. Genet. 53, 1300–1310.

27. Ramasamy, A., Trabzuni, D., Guelfi, S., Varghese, V., Smith, C., Walker, R., De, T., Coin, L., De Silva, R., Cookson, M.R., et al. (2014). Genetic variability in the regulation of gene expression in ten regions of the human brain. Nat. Neurosci. 17, 1418–1428.

28. Ferkingstad, E., Sulem, P., Atlason, B.A., Sveinbjornsson, G., Magnusson, M.I., Styrmisdottir, E.L., Gunnarsdottir, K., Helgason, A., Oddsson, A., Halldorsson, B. V., et al. (2021). Large-scale integration of the plasma proteome with genetics and disease. Nat. Genet. 53, 1712–1721.

29. Folkersen, L., Gustafsson, S., Wang, Q., Hansen, D.H., Hedman, Å.K., Schork, A., Page, K., Zhernakova, D. V., Wu, Y., Peters, J., et al. (2020). Genomic and drug target evaluation of 90 cardiovascular proteins in 30,931 individuals. Nat. Metab. 2, 1135–1148.

30. Weeks, E.M., Ulirsch, J.C., Cheng, N.Y., Trippe, B.L., Fine, R.S., Miao, J., Patwardhan, T.A., Kanai, M., Nasser, J., Fulco, C.P., et al. (2020). Leveraging polygenic enrichments of gene features to predict genes underlying complex traits and diseases. medRxiv.

31. Stelzer, G., Rosen, N., Plaschkes, I., Zimmerman, S., Twik, M., Fishilevich, S., Iny Stein, T., Nudel, R., Lieder, I., Mazor, Y., et al. (2016). The GeneCards suite: From gene data mining to disease genome sequence analyses. Curr. Protoc. Bioinforma. 54, 1.30.1-1.30.33.

32. Ochoa, D., Hercules, A., Carmona, M., Suveges, D., Gonzalez-Uriarte, A., Malangone, C., Miranda, A., Fumis, L., Carvalho-Silva, D., Spitzer, M., et al. (2021). Open Targets Platform: supporting systematic drug–target identification and prioritisation. Nucleic Acids Res. 49, D1302–D1310.

33. Hamosh, A., Scott, A.F., Amberger, J.S., Bocchini, C.A., and McKusick, V.A. (2005). Online Mendelian Inheritance in Man (OMIM), a knowledgebase of human genes and genetic disorders. Nucleic Acids Res. 33, D514–7.

34. Patel, S., Rauf, A., Khan, H., and Abu-Izneid, T. (2017). Renin-angiotensin-aldosterone (RAAS): The ubiquitous system for homeostasis and pathologies. Biomed. Pharmacother. 94, 317–325.

35. Burrell, L.M., Johnston, C.I., Tikellis, C., and Cooper, M.E. (2004). ACE2, a new regulator of the renin–angiotensin system. Trends Endocrinol. Metab. 15, 166.

36. Vickers, C., Hales, P., Kaushik, V., Dick, L., Gavin, J., Tang, J., Godbout, K., Parsons, T., Baronas, E., Hsieh, F., et al. (2002). Hydrolysis of Biological Peptides by Human Angiotensin-converting Enzyme-related Carboxypeptidase. J. Biol. Chem. 277, 14838–14843.

37. Serfozo, P., Wysocki, J., Gulua, G., Schulze, A., Ye, M., Liu, P., Jin, J., Bader, M., Myöhänen, T., García-Horsman, J.A., et al. (2020). Prolyl Oligopeptidase-Dependent Angiotensin II Conversion to Angiotensin-(1-7) in the circulation. Hypertension 75, 173–182.

38. Qiu, S., Pellino, G., Fiorentino, F., Rasheed, S., Darzi, A., Tekkis, P., and Kontovounisios, C. (2017). A Review of the Role of Neurotensin and Its Receptors in Colorectal Cancer. Gastroenterol. Res. Pract. 2017, 6456257.

39. Ngo, D.H., and Vo, T.S. (2019). An Updated Review on Pharmaceutical Properties of Gamma-Aminobutyric Acid. Molecules 24.

40. Liu, X., Hu, C., Bao, M., Li, J., Liu, X., Tan, X., Zhou, Y., Chen, Y., Wu, S., Chen, S., et al. (2016). Genome Wide Association Study Identifies L3MBTL4 as a Novel Susceptibility Gene for Hypertension. Sci. Reports 2016 61 6, 1–11.

41. Hu, C., Zuo, K., Li, K., Gao, Y., Chen, M., Hu, R., Liu, Y., Chi, H., Wang, H., Qin, Y., et al. (2020). p38/JNK Is Required for the Proliferation and Phenotype Changes of Vascular Smooth Muscle Cells Induced by L3MBTL4 in Essential Hypertension. Int. J. Hypertens. 2020, 3123968–3123968.

42. Liu, X.J., Zhang, F.X., Liu, H., Li, K.C., Lu, Y.J., Wu, Q.F., Li, J.Y., Wang, B., Wang, Q., Lin, L.B., et al. (2012). Activin C expressed in nociceptive afferent neurons is required for suppressing inflammatory pain. Brain 135, 391–403.

43. Bygrave, A.M., Sengupta, A., Jackert, E.P., Ahmed, M., Adenuga, B., Nelson, E., Goldschmidt, H.L., Johnson, R.C., Zhong, H., Yeh, F.L., et al. (2021). Btbd11 is an inhibitory interneuron specific synaptic scaffolding protein that supports excitatory synapse structure and function. bioRxiv.

44. Packer, R., Williams, A., Hennah, W., Eisenberg, M., Fawcett, K.A., Pearson, W., Guyatt, A., Edris, A., Hollox, E.J., Rao, B., et al. (2022). Deep-PheWAS: a pipeline for phenotype generation and association analysis for phenome-wide association studies. medRxiv.

45. Bulik-Sullivan, B., Loh, P.R., Finucane, H.K., Ripke, S., Yang, J., Patterson, N., Daly, M.J., Price, A.L., Neale, B.M., Corvin, A., et al. (2015). LD score regression distinguishes confounding from polygenicity in genome-wide association studies. Nat. Genet. 47, 291–295.

46. Bulik-Sullivan, B., Finucane, H.K., Anttila, V., Gusev, A., Day, F.R., Loh, P.R., Duncan, L., Perry, J.R.B., Patterson, N., Robinson, E.B., et al. (2015). An atlas of genetic correlations across human diseases and traits. Nat. Genet. 47, 1236–1241.

47. Hewitt, M.M., Adams, G., Mazzone, S.B., Mori, N., Yu, L., and Canning, B.J. (2016). Pharmacology of Bradykinin-Evoked Coughing in Guinea Pigs. J. Pharmacol. Exp. Ther. 357, 620–8.

48. Canning, B.J. (2010). Afferent nerves regulating the cough reflex: mechanisms and mediators of cough in disease. Otolaryngol. Clin. North Am. 43, 15–vii.

49. Aas, P., and Helle, K.B. (1982). Neurotensin receptors in the rat bronchi. Regul. Pept. 3, 405–413.

50. Djokic, T.D., Dusser, D.J., Borson, D.B., and Nadel, J.A. (1989). Neutral endopeptidase modulates neurotensin-induced airway contraction. J. Appl. Physiol. 66, 2338–2343.

51. Canning, B.J., Mori, N., and Lehmann, A. (2012). Antitussive effects of the peripherally restricted GABAB receptor agonist lesogaberan in guinea pigs: comparison to baclofen and other GABAB receptor-selective agonists. Cough 8, 7.

52. Martvon, L., Kotmanova, Z., Dobrolubov, B., Babalova, L., Simera, M., Veternik, M., Pitts, T., Jakus, J., and Poliacek, I. (2020). Modulation of Cough Reflex by Gaba-Ergic Inhibition in Medullary Raphé of the Cat. Physiol. Res. 69, S151.

53. Dicpinigaitis, P. V., and Dobkin, J.B. (1997). Antitussive effect of the GABA-agonist baclofen. Chest 111, 996–999.

54. Dicpinigaitis, P. V., Dobkin, J.B., Rauf, K., and Aldrich, T.K. (1998). Inhibition of capsaicin-induced cough by the gamma-aminobutyric acid agonist baclofen. J. Clin. Pharmacol. 38, 364–367.

55. Dicpinigaitis, P. V. (1996). Use of baclofen to suppress cough induced by angiotensin-converting enzyme inhibitors. Ann. Pharmacother. 30, 1242–1245.

56. Sutovska, M., Nosalova, G., and Franova, S. (2007). The role of potassium ion channels in cough and other reflexes of the airways. J. Physiol. Pharmacol. 58 Suppl 5, 673–83.

57. Poggioli, R., Benelli, A., Arletti, R., Cavazzuti, E., and Bertolini, A. (1999). Antitussive effect of K+ channel openers. Eur. J. Pharmacol. 371, 39–42.

58. Maranville, J.C., and Cox, N.J. (2016). Pharmacogenomic variants have larger effect sizes than genetic variants associated with other dichotomous complex traits. Pharmacogenomics J. 16, 388–92.

59. Ali, H.A., Lomholt, A.F., Mahmoudpour, S.H., Hermanrud, T., Bygum, A., Von Buchwald, C., Jakobsen, M.A., and Rasmussen, E.R. (2019). Genetic susceptibility to angiotensin-converting enzyme-inhibitor induced angioedema: A systematic review and evaluation of methodological approaches. PLoS One 14, e0224858.

60. Pendergrass, S.A., and Crawford, D.C. (2019). Using Electronic Health Records To Generate Phenotypes For Research. Curr. Protoc. Hum. Genet. 100, e80.

61. Tsang, C., Bottle, A., Majeed, A., and Aylin, P. (2013). Adverse events recorded in English primary care: observational study using the General Practice Research Database. Br. J. Gen. Pract. 63, e534–42.

62. Bradley, S.H., Lawrence, N.R., and Carder, P. (2018). Using primary care data for health research in England -an overview. Futur. Healthc. J. 5, 207–212.

63. Sirugo, G., Williams, S.M., and Tishkoff, S.A. (2019). The Missing Diversity in Human Genetic Studies. Cell 177, 26–31.

64. McCarthy, S., Das, S., Kretzschmar, W., Delaneau, O., Wood, A.R., Teumer, A., Kang, H.M., Fuchsberger, C., Danecek, P., Sharp, K., et al. (2016). A reference panel of 64,976 haplotypes for genotype imputation. Nat. Genet. 48, 1279–1283.

65. Huang, J., Howie, B., McCarthy, S., Memari, Y., Walter, K., Min, J.L., Danecek, P., Malerba, G., Trabetti, E., Zheng, H.F., et al. (2015). Improved imputation of low-frequency and rare variants using the UK10K haplotype reference panel. Nat. Commun. 6, 1–9.

66. The 1000 Genomes Project Consortium (2015). A global reference for human genetic variation. Nature 526, 68–74.

67. Taliun, D. (2021). Sequencing of 53,831 diverse genomes from the NHLBI TOPMed Program. Nature 590, 290–299.

68. Shrine, N., Guyatt, A.L., Erzurumluoglu, A.M., Jackson, V.E., Hobbs, B.D., Melbourne, C.A., Batini, C., Fawcett, K.A., Song, K., Sakornsakolpat, P., et al. (2019). New genetic signals for lung function highlight pathways and chronic obstructive pulmonary disease associations across multiple ancestries. Nat. Genet. 51, 481–493.

69. Mbatchou, J., Barnard, L., Backman, J., Marcketta, A., Kosmicki, J.A., Ziyatdinov, A., Benner, C., O’Dushlaine, C., Barber, M., Boutkov, B., et al. (2021). Computationally efficient whole-genome regression for quantitative and binary traits. Nat. Genet. 53, 1097–1103.

70. Chang, C.C., Chow, C.C., Tellier, L.C.A.M., Vattikuti, S., Purcell, S.M., and Lee, J.J. (2015). Second-generation PLINK: Rising to the challenge of larger and richer datasets. Gigascience 4, 7.

71. Willer, C.J., Li, Y., and Abecasis, G.R. (2010). METAL: Fast and efficient meta-analysis of genomewide association scans. Bioinformatics 26, 2190–2191.

72. D. Turner S. (2018). qqman: an R package for visualizing GWAS results using Q-Q and manhattan plots. J. Open Source Softw. 3, 731.

73. Yang, J., Ferreira, T., Morris, A.P., Medland, S.E., Madden, P.A.F., Heath, A.C., Martin, N.G., Montgomery, G.W., Weedon, M.N., Loos, R.J., et al. (2012). Conditional and joint multiple-SNP analysis of GWAS summary statistics identifies additional variants influencing complex traits. Nat. Genet. 44, 369–375.

74. Yang, J., Lee, S.H., Goddard, M.E., and Visscher, P.M. (2011). GCTA: A tool for genome-wide complex trait analysis. Am. J. Hum. Genet. 88, 76–82.

75. Weissbrod, O., Hormozdiari, F., Benner, C., Cui, R., Ulirsch, J., Gazal, S., Schoech, A.P., van de Geijn, B., Reshef, Y., Márquez-Luna, C., et al. (2020). Functionally informed fine-mapping and polygenic localization of complex trait heritability. Nat. Genet. 52, 1355–1363.

76. Wang, G., Sarkar, A., Carbonetto, P., and Stephens, M. (2020). A simple new approach to variable selection in regression, with application to genetic fine mapping. J. R. Stat. Soc. Ser. B Stat. Methodol. 82, 1273–1300.

77. Gazal, S., Finucane, H.K., Furlotte, N.A., Loh, P.R., Palamara, P.F., Liu, X., Schoech, A., Bulik-Sullivan, B., Neale, B.M., Gusev, A., et al. (2017). Linkage disequilibrium-dependent architecture of human complex traits shows action of negative selection. Nat. Genet. 49, 1421–1427.

78. Gazal, S., Loh, P.R., Finucane, H.K., Ganna, A., Schoech, A., Sunyaev, S., and Price, A.L. (2018). Functional architecture of low-frequency variants highlights strength of negative selection across coding and non-coding annotations. Nat. Genet. 50, 1600–1607.

79. Gazal, S., Marquez-Luna, C., Finucane, H.K., and Price, A.L. (2019). Reconciling S-LDSC and LDAK functional enrichment estimates. Nat. Genet. 51, 1202–1204.

80. Wakefield, J. (2009). Bayes factors for Genome-wide association studies: Comparison with P-values. Genet. Epidemiol. 33, 79–86.

81. Wang, K., Li, M., and Hakonarson, H. (2010). ANNOVAR: Functional annotation of genetic variants from high-throughput sequencing data. Nucleic Acids Res. 38, e164.

82. Kircher, M., Witten, D.M., Jain, P., O’Roak, B.J., Cooper, G.M., and Shendure, J. (2014). A general framework for estimating the relative pathogenicity of human genetic variants. Nat. Gene 46, 310–315.

83. McLaren, W., Gil, L., Hunt, S.E., Riat, H.S., Ritchie, G.R.S., Thormann, A., Flicek, P., and Cunningham, F. (2016). The Ensembl Variant Effect Predictor. Genome Biol. 17, 122.

84. Watanabe, K., Taskesen, E., Van Bochoven, A., and Posthuma, D. (2017). Functional mapping and annotation of genetic associations with FUMA. Nat. Commun. 8, 1–10.

85. Ghoussaini, M., Mountjoy, E., Carmona, M., Peat, G., Schmidt, E.M., Hercules, A., Fumis, L., Miranda, A., Carvalho-Silva, D., Buniello, A., et al. (2021). Open Targets Genetics: systematic identification of trait-associated genes using large-scale genetics and functional genomics. Nucleic Acids Res. 49, D1311–D1320.

86. MacArthur, J., Bowler, E., Cerezo, M., Gil, L., Hall, P., Hastings, E., Junkins, H., McMahon, A., Milano, A., Morales, J., et al. (2017). The new NHGRI-EBI Catalog of published genome-wide association studies (GWAS Catalog). Nucleic Acids Res. 45, D896–D901.

87. Neale Lab (2018). UK Biobank GWAS Round 2. http://www.nealelab.is/uk-biobank/.

88. Lee Lab (2018). UK-Biobank Single Variant Association Analysis Results (Binary Phenome Analysis). https://www.leelabsg.org/resources.

