## Supplementary Figures for "Genome-wide association study of ACE inhibitor-induced cough implicates neuropeptides and shows genetic overlap with chronic dry cough"

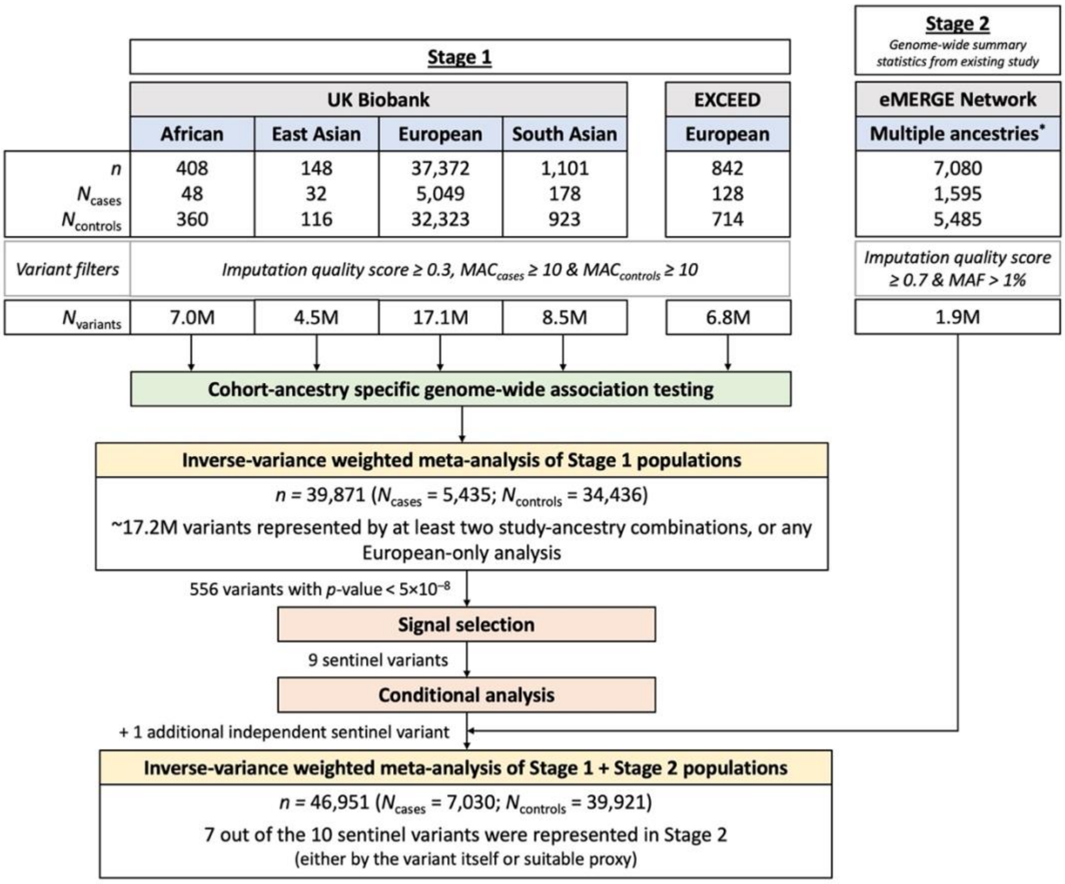


**Supplementary Figure 1** Overview of study design.
*Genetic ancestry groups defined as 'European', 'African' and 'Other' by study authors[1].

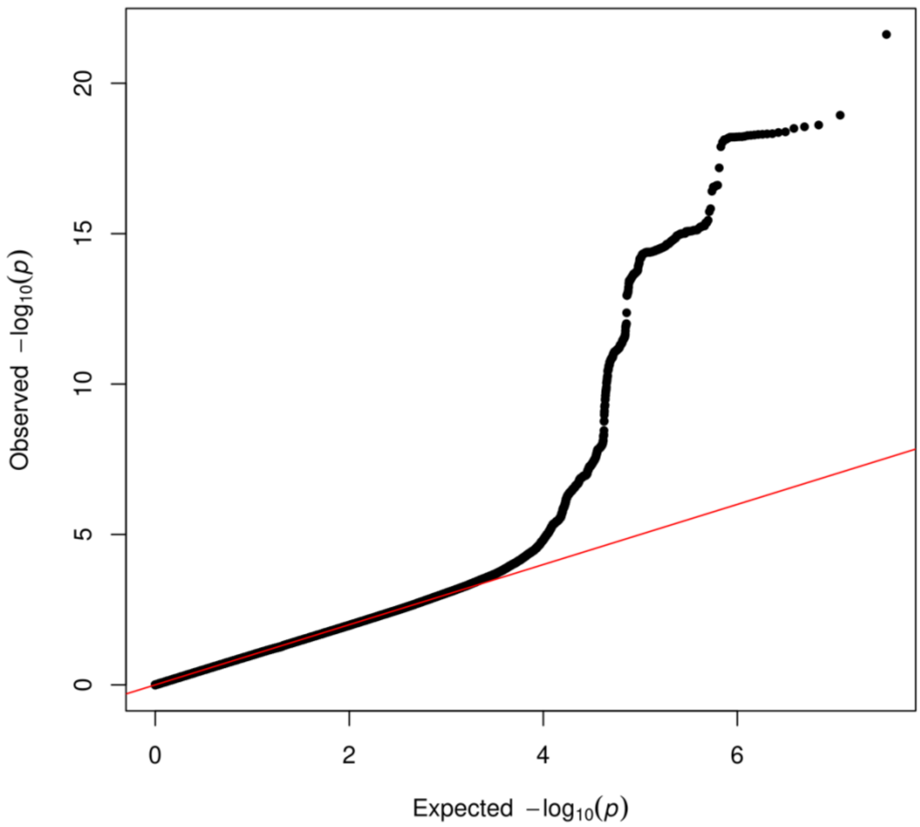


**Supplementary Figure 2** Quantile-quantile plot for Stage 1 genome-wide meta-analysis of ACEI induced cough.

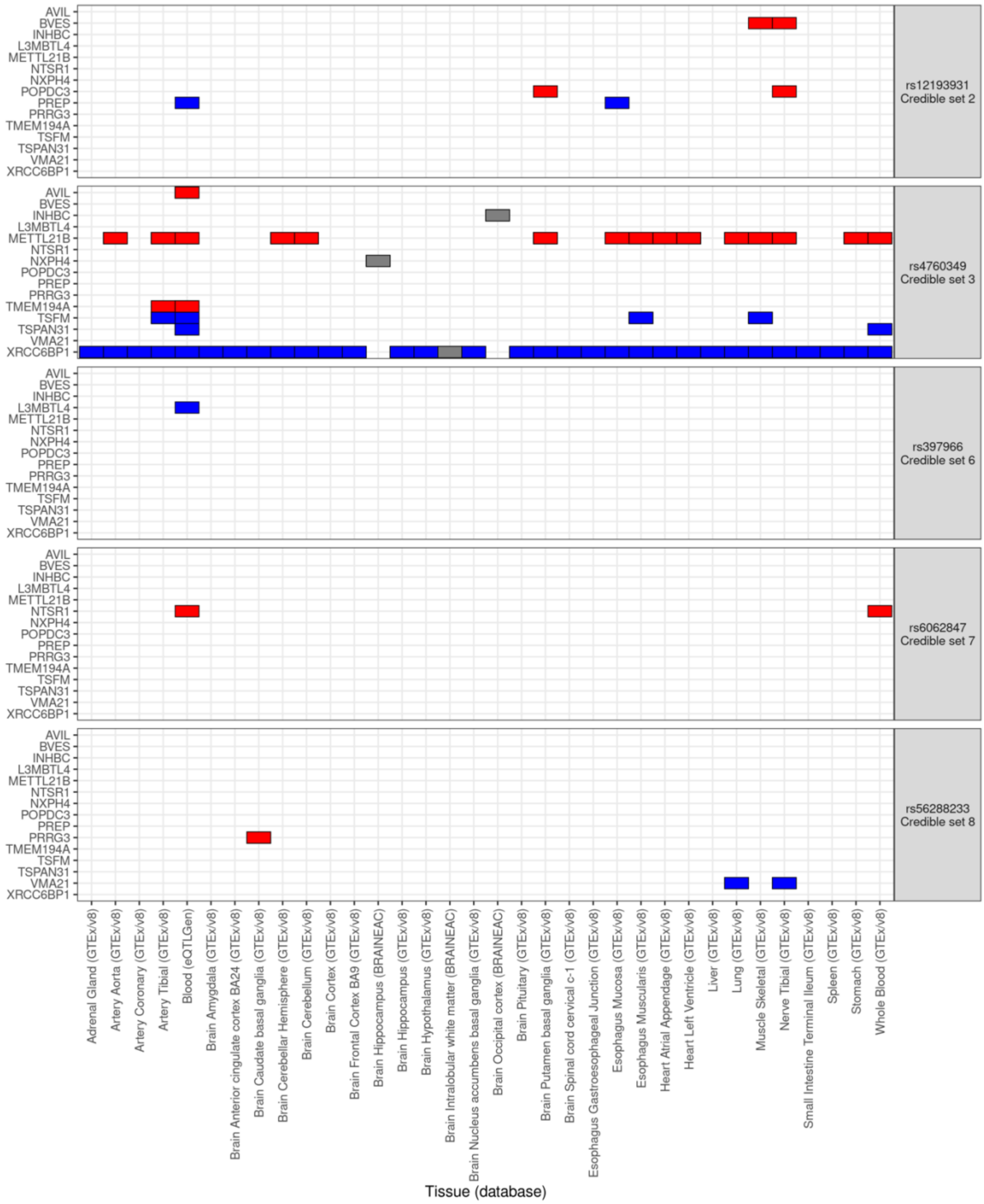


**Supplementary Figure 3** Summary of 15 genes for which expression is significantly (FDR <0.05) affected by variants in refined credible sets. Credible set variants without significant eQTL results have been omitted. Red tiles denote a significant increase in expression with respect to increasing risk of ACEI-induced cough. Blue tiles denote a significant decrease in expression with respect to increasing risk of ACEI-induced cough. Grey tiles denote when effect direction could not be predicted in BRAINEAC dataset, given that the tested allele was not provided. Facet labelling describes sentinel variant and corresponding credible set number.

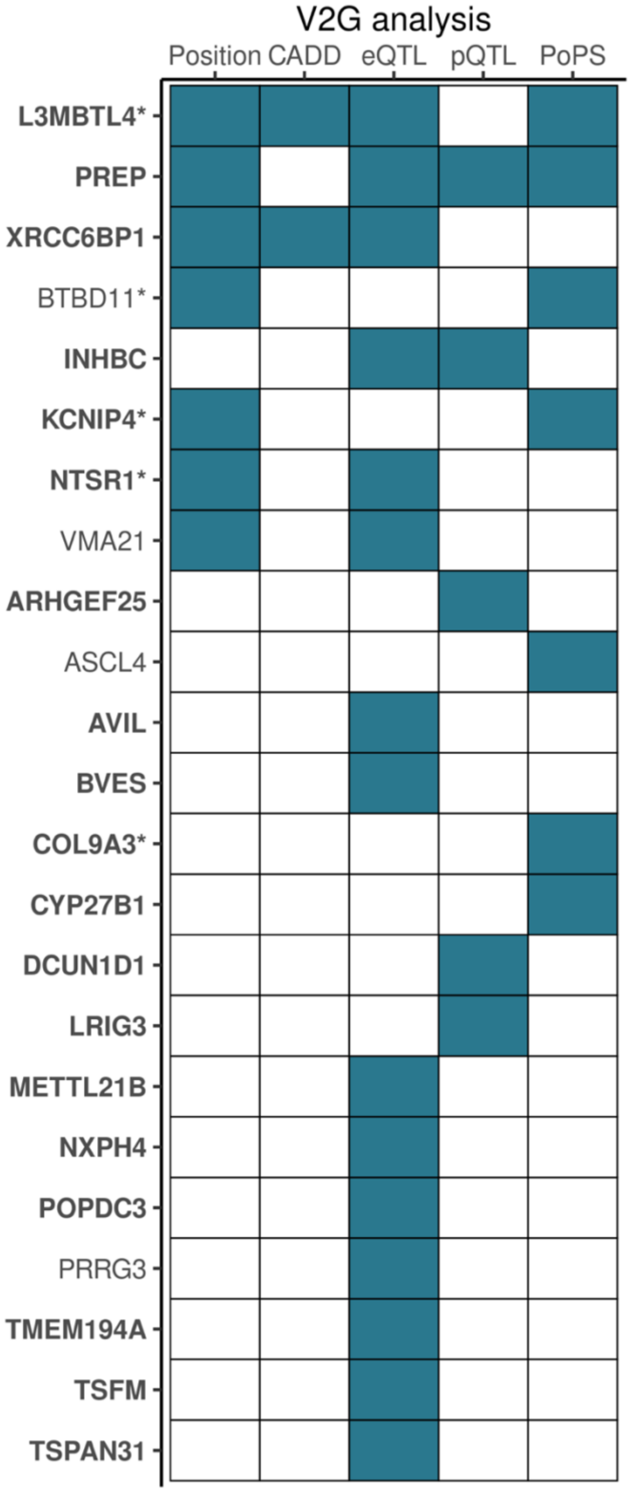


**Supplementary Figure 4** Genes identified by each variant-to-gene (V2G) mapping analysis based on 95% credible set variants with PIP >10% (refined credible set). Filled tiles denote gene identified by specific analysis. Genes identified by at least one refined credible set variant from fine-mapping of genome-wide significant loci in joint meta-analysis are highlighted bold, and nonbold if the fine-mapped locus was genome-wide significant in Stage 1 only (without representation in Stage 2).
*Genes identified by at least one V2G mapping analysis with a refined credible set variant with PIP >50%.
Abbreviations: CADD, Combined Annotation Dependent Depletion; eQTL, expression quantitative trait loci; PoPS, Polygenic Priority Score; pQTL, protein quantitative trait loci.

A

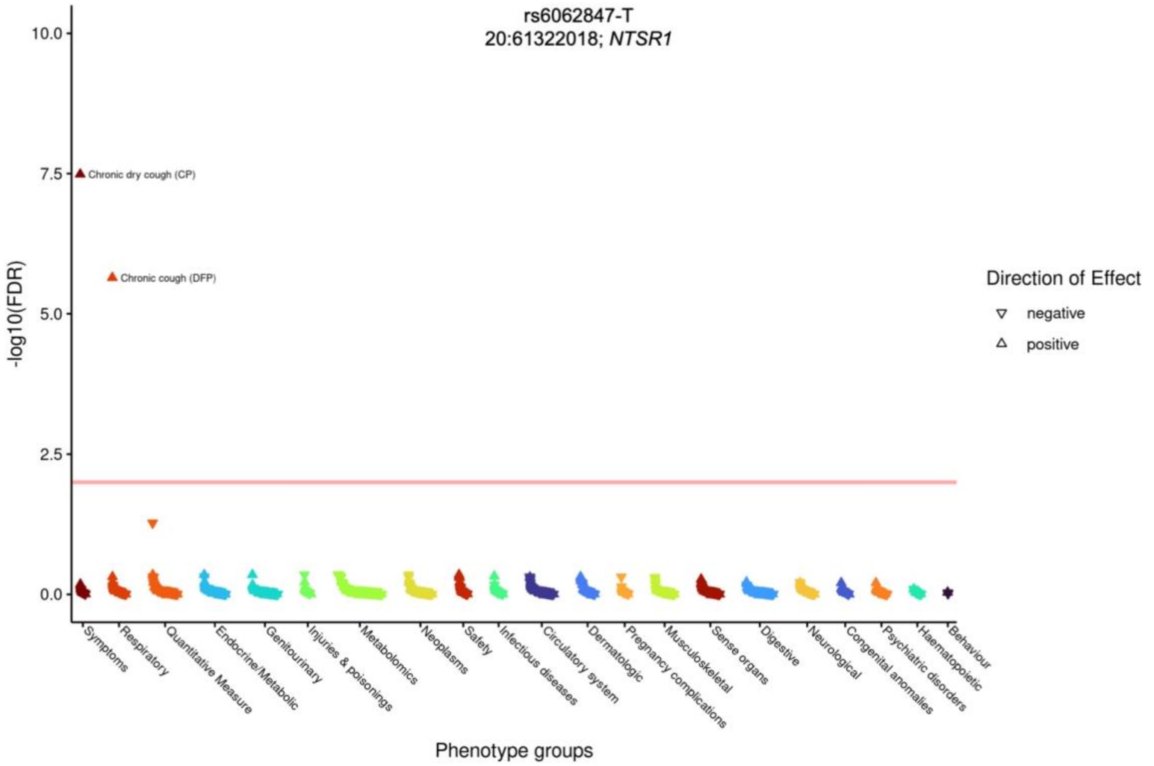

rs6062847-T
20:61322018; *NTSR1*
-log10(FDR)
10.0
7.5
5.0
2.5
0.0
Chronic dry cough (CP)
Chronic cough (DFP)
Direction of Effect
negative
positive
Symptoms
Respiratory
Quantitative Measure
Endocrine/Metabolic
Genitourinary
Injuries & poisonings
Metabolomics
Neoplasms
Safety
Infectious diseases
Circulatory system
Dermatologic
Pregnancy complications
Musculoskeletal
Sense organs
Digestive
Neurological
Congenital anomalies
Psychiatric disorders
Haematopoietic
Behaviour
Phenotype groups


B

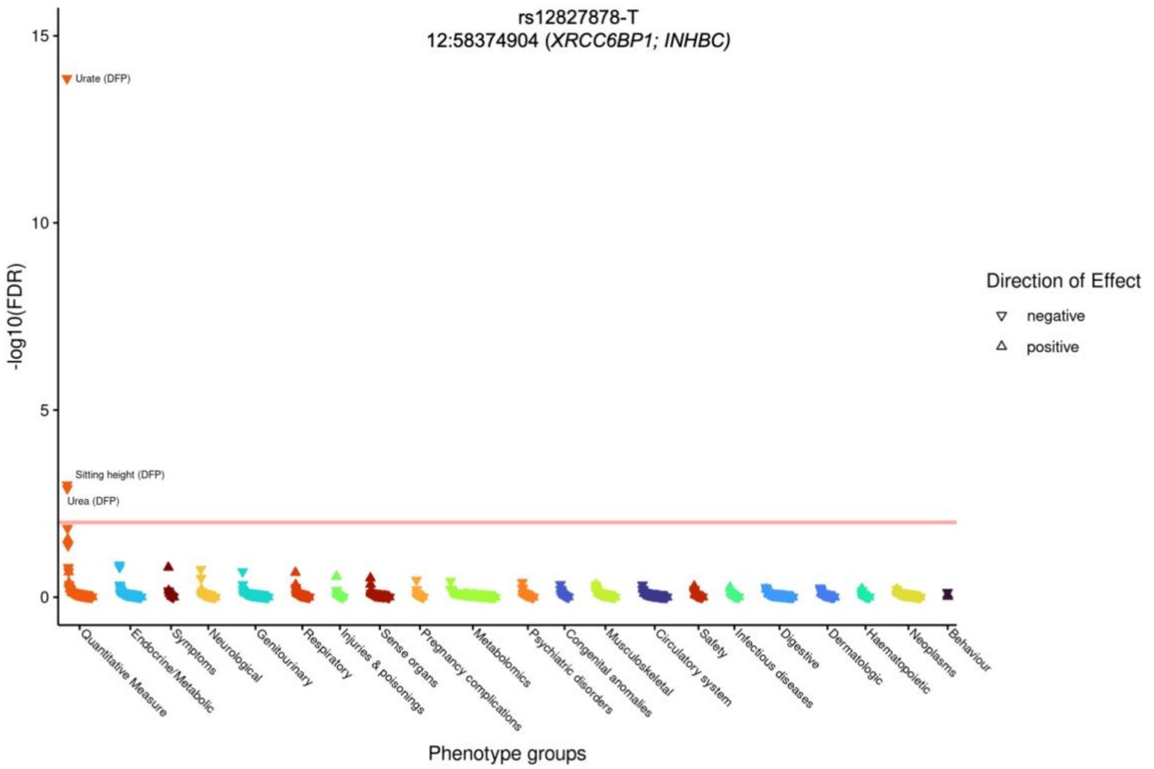

rs12827878-T
12:58374904 (*XRCC6BP1; INHBC)*
-log10(FDR)
15
10
5
0
Urate (DFP)
Sitting height (DFP)
Urea (DFP)
Direction of Effect
negative
positive
Quantitative Measure
Endocrine/Metabolic
Symptoms
Neurological
Genitourinary
Respiratory
Injuries & poisonings
Sense organs
Pregnancy complications
Metabolomics
Psychiatric disorders
Congenital anomalies
Musculoskeletal
Circulatory system
Safety
Infectious diseases
Digestive
Dermatologic
Haematopoietic
Neoplasms
Behaviour
Phenotype groups

C

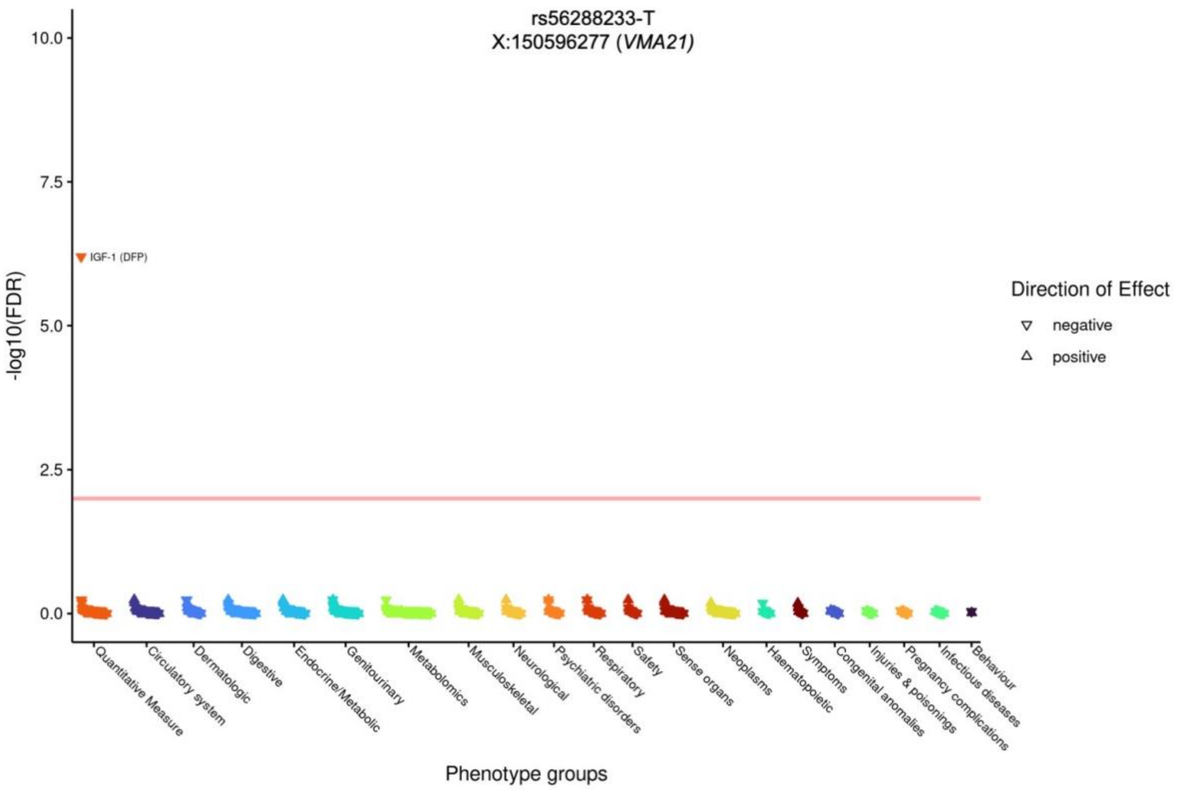


**Supplementary Figure 5** Phenome-wide association study (PheWAS) results for (A) rs6062847-T, (B) rs12827878-T and (C) rs56288233-T on up to 1,913 UK Biobank-derived traits and diseases. Each panel is titled with variant identifier and allele which confers increasing risk of ACEI induced cough, genomic coordinates (build hg19) and putative causal gene(s). The red horizontal line represents the phenome-wide significance threshold (false discovery rate [FDR] of 0.01).
Abbreviations: CP, composite phenotype; DFP, data-field phenotype.

References

1. Mosley, J. D. *et al.* A genome-wide association study identifies variants in KCNIP4 associated with ACE inhibitor-induced cough. *Pharmacogenomics J.* **16**, 231–237 (2016).
